# Identification of Plasma Protein Biomarkers for Early-Onset Stroke through Mendelian Randomization

**DOI:** 10.64898/2026.09.14.26363028

**Authors:** Anna L. Huynh, Brady J. Gaynor, John W. Cole, Patrick F. McArdle, Huichun Xu, Christina Jern, Guillaume Pare, Steven J. Kittner, Michael Chong, Tara M. Stanne, Braxton D. Mitchell, the Early-Onset Stroke Consortia

**Affiliations:** University of Maryland, College Park, Maryland, United States; Department of Medicine, University of Maryland School of Medicine, Baltimore, MD, United States; Departments of Neurology, Baltimore VAMC and University of Maryland School of Medicine, Baltimore, Maryland, United States; Department of Medicine, School of Medicine, University of Maryland Baltimore, Baltimore, MD, United States; Institute of Biomedicine, Department of Laboratory Medicine, Sahlgrenska Academy, University of Gothenburg, Gothenburg, Sweden and Department of Clinical Genetics and Genomics, Sahlgrenska University Hospital, Region Västra Götaland Gothenburg, Sweden; Department of Pathology and Molecular Medicine, Faculty of Health Sciences, McMaster University, Hamilton, Ontario, Canada; Department of Neurology, University of Maryland School of Medicine, Baltimore, Maryland, United States; Department of Pathology and Molecular Medicine, Michael G. DeGroote School of Medicine, Hamilton, Ontario, Canada; Department of Medicine, University of Maryland School of Medicine, Baltimore, Maryland, United States

**Keywords:** Ischemic Stroke, Mendelian Randomization, Risk Factors, Young Adults, Proteins

## Abstract

**Background:** Stroke incidence is declining in older adults but rising in younger populations. Circulating plasma proteins are promising biomarkers of vascular disease, yet their causal role in early-onset stroke (EOS) remains unclear. We used proteome-wide Mendelian randomization (MR) to identify proteins causally associated with EOS and its subtypes.

**Methods:** We conducted two-sample MR using protein quantitative trait loci from 54,219 UK Biobank participants and summary-level association results from up to 11,114 EOS cases (onset <60 years) and 435,540 controls in the Early-Onset Stroke Consortium. Variants within ±200 kb of protein-coding genes associated with protein levels (p ≤1×10⁻⁵) were clumped for linkage disequilibrium and used as instrumental variables. Causal effects were estimated using inverse-variance weighted MR, with sensitivity analyses to assess robustness. Stroke subtypes were defined using TOAST criteria.

**Results:** Among 1,041 proteins tested, 11 were significantly associated with EOS after multiple testing correction. Genetically predicted higher levels of proteins encoded by *ABO*, *F11*, *F7*, *MLN*, *BTD*, *MEP1B*, and *VAMP8* increased EOS risk (ORs ∼1.05-1.32 per SD), whereas higher levels of proteins encoded by *FN1*, *GRK5*, *EHBP1*, and *FCN2* were protective. Eight additional proteins showed subtype-specific associations. Key signals converged on coagulation and thromboinflammatory pathways, particularly *ABO*, *F11*, and *F7*, with consistent evidence across analyses. Cross-trait analyses demonstrated overlap with late-onset stroke, venous thromboembolism, and cardiometabolic risk factors, supporting shared biological mechanisms. Drug-target mapping highlighted Factor XI as a leading therapeutic target, supported by existing inhibitors (e.g., abelacimab, milvexian).

**Conclusions:** This large-scale proteome-wide MR study identified multiple plasma proteins with likely causal roles in EOS, implicating coagulation, platelet activation, extracellular matrix remodeling, and immune pathways in disease etiology. These findings highlight biologically plausible biomarkers and prioritize druggable targets, particularly Factor XI, for prevention of EOS and demonstrate the value of proteogenomic approaches for uncovering disease mechanisms and informing precision prevention strategies.

**Brief Summary:** Proteome-wide MR of 1,041 plasma proteins identified causal drivers of early-onset stroke, with strongest effects in coagulation (ABO, F7, F11) and immune-inflammatory pathways.

## Introduction

Stroke is a leading cause of death and disability, with more than 795,000 new and recurrent cases annually in the United States as of 2025.^1^ Although most strokes occur in individuals over the age of 50 years, ≈10% of strokes occur in younger adults aged 18 to 50 years.^2^ Since the year 2000, incidence rates of ischemic stroke in high-income countries have been declining among older individuals but have been increasing in individuals aged <55 years.^3^ presenting unique clinical and public health challenges.

Identifying biomarkers for incident stroke is a high priority because they could shed light on underlying pathophysiological processes and help identify individuals at high risk of stroke or recurrent stroke. Proteins are particularly appealing as biomarkers due to their potential links to susceptibility genes. Recent advances in high-throughput proteomic technologies have enabled comprehensive, accurate, and cost-effective assessments of multiple proteins in large-scale studies. Epidemiologic research has reported associations between circulating levels of several proteins and incident stroke ^4–10^, but these studies have generally been limited by small numbers of incident events, resulting in low statistical power and leaving observed associations vulnerable to common limitations of observational studies, such as confounding.

As a complement to epidemiologic studies, Mendelian randomization (MR) has emerged as a powerful approach for inferring causal relationships between exposures and outcomes by using genetic variants as instrumental variables. Mendelian randomization is particularly well suited for estimating causal associations of proteins on disease outcomes because the genes encoding proteins are known, allowing genetic instruments to be tied specifically and exclusively to the exposure. Recent Mendelian randomization studies have now reported causal associations between multiple proteins and ischemic stroke.^11–24^

In this study, we conducted two-sample Mendelian randomization analyses using genetic instruments for over 1,000 plasma proteins to identify novel causal determinants of early onset stroke (EOS) and its major subtypes. Previous Mendelian randomization studies of proteins and stroke have primarily focused on strokes occurring in individuals over age 60. In contrast, our study targets early onset stroke (EOS), that is, stroke before the age of 60, where heritability is higher ^25–27^ and the etiological subtype distribution differs compared to ischemic stroke at older age.

By integrating summary genetic association results for proteomics from the UK Biobank and for EOS from the Early-Onset Stroke Consortium, we aimed to elucidate biological pathways underlying stroke susceptibility in younger individuals and identify potential therapeutic targets for stroke prevention.

## Methods

### Study design

This study was performed according to the Strengthening the Reporting of Observational Studies in Epidemiology Using Mendelian Randomization (STROBE-MR) and the recommended guidelines.^28^ We employed a 2-sample Mendelian randomization design using IVW as our primary method of analysis to assess the causal estimates of plasma protein levels on early onset ischemic stroke. Importantly, no plasma protein measurements were obtained from EOS cases included in the outcome dataset. Protein levels were genetically predicted using pQTLs derived from the UK Biobank proteomic GWAS. Consequently, MR estimates reflect the effects of lifelong genetically determined differences in protein levels rather than protein concentrations measured following stroke onset. This distinction minimizes concerns regarding reverse causation and confounding by acute stroke-related factors.

### Sources of data

Exposure date (protein quantitative loci, or pQTLs) were obtained from published GWAS summary statistics of 54,219 individuals of predominately (95%) European ancestry (mean age 56.8 yrs) from the UK Biobank–Pharma Proteomics Project (UKB-PPP).^19^ Plasma protein levels were measured using the antibody-based OLINK Explore 3072 platform. For this study we analyzed all proteins from the Explore 3072 inflammation and cardiometabolic panels (n = 1,540).

Outcome data were derived from genome-wide association statistics of 11,114 early-onset stroke cases and 435,540 controls of European ancestry from the Early-Onset Stroke Consortium (EOSC).^29^ The EOSC included ischemic stroke cases ages 18-59 years, which was established a priori in the original consortium genome-wide association study and has been used in subsequent genetic investigations of early-onset ischemic stroke. All stroke outcomes evaluated in this study were ischemic strokes. The UK Biobank dataset contributed only protein quantitative trait loci (pQTL) information and did not contribute stroke cases. Stroke cases in EOSC underwent brain imaging at each participating site to confirm ischemic stroke diagnosis, exclude alternate diagnoses, and support subtype classification. Additional screening was performed in some, but not all, contributing studies to exclude cases attributable to known monogenic causes (e.g., sickle cell disease) or non-genetic causes (e.g., drug use, procedural complications). Ischemic stroke subtyping was performed using the Trial of ORG 10172 in Acute Stroke Treatment (TOAST) criteria ^30^ in most, but not all, cohorts; subtype-specific case counts are provided in **Table S1**. TOAST subtype assignments were generated within participating EOSC studies according to established TOAST protocols. Because EOSC represents a consortium of multiple studies, diagnostic evaluations varied somewhat across contributing cohorts but generally included neuroimaging and clinical evaluation sufficient for ischemic stroke and subtype classification. Additional details regarding diagnostic work-up and adjudication procedures are available in the original EOSC publication. ^29^

Genotypes were aligned to the GRCh38 (hg38) reference genome and imputed using the TOPMed reference panel via the University of Michigan Imputation Server.^31^

### Instrument selection

We used cis-acting genetic variants in protein-coding genes as instrumental variables for circulating protein levels. For each gene, we extracted all variants within ± 200kb of gene boundaries from the corresponding pQTL summary statistics. Variants were retained if they were associated with protein levels at p ≤ 1×10⁻⁵ and had minor allele frequency > 0.01.

SNP identifiers were converted to GRCh38 genomic coordinates and harmonized with the outcome dataset. Variants were then pruned for linkage disequilibrium using PRSice,^32^ applying a 250 kb window and r² threshold < 0.1.

### Mendelian Randomization Analysis

We estimated the causal effect of protein levels on EOS risk using two-sample MR in R with the MendelianRandomization package.^33^ The primary analysis used the IVW method. To account for multiple hypotheses testing, statistical significance was determined using a Bonferroni-corrected thresholds (p< 0.05 divided by the number of proteins tested). Effect estimates are reported as odds ratios (OR) corresponding to a one standard deviation (SD) increase in genetically predicted protein levels.

The validity of MR estimates relies on three core instrumental variable assumptions: (1) the genetic variants are associated with the exposure (relevance), (2) the variants are independent of confounders (independence), and (3) the variants influence the outcome only through the exposure (exclusion restriction). Sensitivity analyses included weighted median and MR-Egger regression approaches, including evaluation of the MR-Egger intercept.^34^ Heterogeneity in variant-specific effect sizes was assessed using Cochran Q-statistic and MR-PRESSO.

### Association of EOS-associated proteins with related vascular phenotypes

We further evaluated whether proteins associated with EOS exhibited casual effects on related vascular outcomes, including late onset stroke, LOS; venous thromboembolism, VTE; and established stroke risk factors (i.e., hypertension; type 2 diabetes mellitus, TD2; LDL cholesterol levels; and body mass index, BMI).

Two-sample MR analyses were conducted using summary statistics restricted to individuals of European ancestry to match the proteomic GWAS population. These datasets included 9,272 LOS cases (stroke onset age ≥ 60 years), 81,190 VTE cases, 320,429 hypertension cases, and 74,124 T2D cases. Summary statistics for LDL cholesterol and BMI were based on 1,320,016 and approximately 700,000 individuals, respectively (**Table S2**).

For proteins associated with EOS, we tested whether their causal effects differed from those observed for LOS by comparing effect estimates using a Wald test. Specifically, we calculated a test statistic based on the difference between the beta estimates divided by the standard error of the difference (derived from the sum of the squared standard errors). We accounted for multiple testing using a Bonferroni correction based on the number of significantly associated proteins evaluated (p = 0.05 / 19 = 0.0026).

### MR analysis of proteins previously associated epidemiologically with incident stroke risk

To further investigate the relationship between circulating proteins and EOS, we systematically identified proteins reported in six published prospective cohort studies that evaluated baseline plasma or serum protein levels in relation to incident ischemic stroke,.^4,6,7,10,13,35^ These studies, conducted in populations for which the incident stroke cases were dominated primarily by late-onset cases, provide observational evidence linking specific proteins to stroke incidence. We then used MR to assess whether these proteins were causally associated with EOS using MR.

#### Drug targets

To identify potential therapeutic targets that could be repurposed for disease treatment, we queried the DGIdb database (https://www.dgidb.org/), which catalogs drug–gene interactions and druggable genes from a wide range of sources, including curated databases and published literature.^36^ In DGIdb, drug–gene relationships are systematically identified, normalized, and grouped through an ontology-based harmonization pipeline that integrates data from diverse drug resources, consolidating all known synonyms and identifiers for a therapeutic agent under a unified drug concept. This framework enables accurate and comprehensive mapping between EOS-associated genes and known or potential drug interactions. To enhance robustness, we retained only drug-gene interactions with an interaction score > 0.50.

#### Independent data access and analysis

Ms. Hyunh, Mr. Gaynor, and Drs. Xu and Mitchell had full access to all data in the study. Drs. Xu and Mitchell take responsibility for study integrity and the data analysis.

#### Data availability

The two sample MR analyses presented in this study are based on published summary statistics. Summary level GWAS statistics from EOSC are available through the Cerebrovascular Disease Genetics Portal. The sources of summary statistics for the other outcomes are provided in Table S2.

## Results

Of the 1,540 inflammation and cardiometabolic proteins analyzed, we constructed genetic instruments for 1,041 proteins with at least three or more independently associated pQTLs. The instruments contained between 3 and 132 variants, with the number of variants included in each instrument reported in **Table S3**. The F values, a measure of the strength of the instrumental variable, exceeded 23 for all included variants.

Because the proteomic analyses were conducted using summary-level genetic association statistics rather than individual participant data, detailed clinical characteristics, health history and medication use relative to timing of blood collection were not available for analysis within the present study.

At a Bonferroni significance threshold of p < 4.80E-05 (0.05/1041 proteins tested), we identified 11 proteins as causally associated with EOS and 8 proteins associated with an EOS subtype. The 19 associated proteins and the IVW estimates for these causal associations are shown in **Table 1**. The subtype-specific associations included *MMP10* with large artery atherosclerotic stroke, ITGA6/TREH/CPXM2/LGALS3 with small artery occlusion, and GRK5/GRAP2/IL22RA1 with cardioembolic stroke, supporting the existence of distinct biologic mechanisms across etiologic stroke subtypes. Sensitivity analyses provided little evidence for pleiotropy or confounding. Specifically, the weighted median and MR Egger estimates were consistent with the IVW estimates, and none of the MR-Egger intercepts deviated significantly from zero, indicating no evidence of directional pleiotropy (**Table S4**).

**Table 1.** IVW estimates of causal associations of proteins with EOS and subtypes

| Protein |  | Phenotype | # SNPs (EOS) | OR | CI_L | CI-U | p-val |
| --- | --- | --- | --- | --- | --- | --- | --- |
| ABO |  | All EOS | 58 | 1.111 | 1.087 | 1.135 | 1.34E-21 |
| F11 |  | All EOS | 62 | 1.191 | 1.134 | 1.251 | 2.56E-12 |
| FN1 |  | All EOS | 43 | 0.854 | 0.808 | 0.902 | 1.67E-08 |
| GRK5 |  | All EOS | 28 | 0.783 | 0.719 | 0.854 | 2.93E-08 |
| MLN |  | All EOS | 63 | 1.130 | 1.080 | 1.182 | 1.13E-07 |
| F7 |  | All EOS | 45 | 1.111 | 1.064 | 1.160 | 1.82E-06 |
| BTD |  | All EOS | 42 | 1.083 | 1.044 | 1.124 | 2.55E-05 |
| MEP1B |  | All EOS | 55 | 1.047 | 1.025 | 1.070 | 2.89E-05 |
| EHBP1 |  | All EOS | 17 | 0.754 | 0.659 | 0.861 | 3.16E-05 |
| FCN2 |  | All EOS | 63 | 0.929 | 0.896 | 0.962 | 3.94E-05 |
| VAMP8 |  | All EOS | 19 | 1.317 | 1.155 | 1.501 | 4.05E-05 |
| LYVE1 |  | All EOS | 20 | 1.324 | 1.157 | 1.516 | 4.65E-05 |
| MMP10 |  | EOS-LAA | 53 | 0.739 | 0.640 | 0.855 | 4.48E-05 |
| TREH |  | EOS - SAO | 32 | 0.813 | 0.744 | 0.888 | 4.22E-06 |
| ITGA6 |  | EOS - SAO | 36 | 2.273 | 1.585 | 3.260 | 8.12E-06 |
| CPXM2 |  | EOS - SAO | 37 | 1.833 | 1.385 | 2.426 | 2.26E-05 |
| LGALS3 |  | EOS - SAO | 27 | 0.688 | 0.576 | 0.822 | 3.96E-05 |
| GRK5 |  | EOS - CE | 28 | 0.438 | 0.335 | 0.573 | 1.72E-09 |
| GRAP2 |  | EOS - CE | 3 | 13.014 | 3.983 | 42.514 | 2.15E-05 |
| IL22RA1 |  | EOS - CE | 24 | 0.554 | 0.420 | 0.730 | 2.77E-05 |

Significance threshold set at $p < 0.05/1041$ , or $p < 4.80E-05$ (see text)

MR PRESSO and Cochran’s Q heterogeneity tests were subsequently performed. Seventeen of these proteins showed no evidence of heterogeneity among SNP effects, although modest evidence for heterogeneity (0.01<p<0.05) was observed for *FCN2* (all EOS) and *LGALS3* (EOS-SAO). Subsequent leave-one-out sensitivity analyses demonstrated consistent causal estimates for these proteins after sequential exclusion of individual *FCN2* and *LGALS3* instruments, indicating that the observed inverse association between genetically predicted levels of these proteins and stroke risk was not driven by any single genetic variant and the identified associations were robust despite modest heterogeneity.

### Associations with LOS

Of the 19 proteins causally associated with EOS or one of its subtypes, three were also significantly associated with LOS (coagulation factor XI (*F11*), G protein-coupled receptor kinase 5 (*GRK5)*, and coagulation factor VII *(F7*)) at the Bonferroni-corrected threshold for significance (0.05/19 = 0.0026) (**Table 2**). Heterogeneity tests indicated that the effect sizes for four of the proteins (*ABO*, *FN1*, *ITGA6*, and *EHBP1*) differed significantly between early and late onset stroke (**Table S5**). In each case, effect sizes were larger in EOS than in LOS.

**Table 2.** 19 EOS-associated proteins and their causal associations with LOS, VTE, and stroke-related risk factors

| Protein | Gene | Associated Phenotype |  | EOS | LOS | VTE-EUR | HBP | BMI | T2D | LDL-C | Potential mechanistic pathways |
| --- | --- | --- | --- | --- | --- | --- | --- | --- | --- | --- | --- |
| ABO glycosyltransferase | ABO | All EOS |  | ↑ |  | ↑ |  |  | ↑ | ↑ | prothrombotic |
| Coagulation factor XI | F11 | All EOS |  | ↑ | ↑ | ↑ |  | ↑ |  |  | coagulation/platelet activation |
| G protein-coupled receptor kinase 5 | GRK5 | EOS - CE |  | ↓ | ↓ | ↓ |  |  |  |  | platelet secretion and thrombin-PAR signaling |
| Fibronectin | FN1 | All EOS |  | ↓ |  |  |  |  |  | ↑ | atherosclerotic matrix remodeling |
| G protein-coupled receptor kinase 5 | GRK5 | All EOS |  | ↓ |  |  |  |  |  |  | platelet secretion and thrombin-PAR signaling |
| Motilin | MLN | All EOS |  | ↑ |  |  | ↓ |  |  |  |  |
| Coagulation factor VII | F7 | All EOS |  | ↑ | ↑ |  |  |  |  |  | coagulation/platelet activation |
| Trehalase | TREH | EOS - SAO |  | ↓ |  |  |  |  |  |  |  |
| Integrin alpha-6 | ITGA6 | EOS - SAO |  | ↑ |  |  |  |  |  |  |  |
| GRB2-related adaptor protein 2 | GRAP2 | EOS - CE |  | ↑ |  |  |  |  |  |  |  |
| Carboxypeptidase X2 | CPXM2 | EOS - SAO |  | ↑ |  |  |  |  |  |  |  |
| Biotinidase | BTD | All EOS |  | ↑ |  |  |  |  |  |  |  |
| Interleukin-22 receptor subunit alpha-1 | IL22RA1 | EOS - CE |  | ↓ |  |  |  |  |  |  |  |
| Meprin B | MEP1B | All EOS |  | ↑ |  |  | ↓ |  | ↓ |  |  |
| EH domain-binding protein 1 | EHBP1 | All EOS |  | ↓ |  |  |  |  |  |  |  |
| Ficolin-2 | FCN2 | All EOS |  | ↓ |  |  |  |  |  | ↓ | lectin-complement activation |
| Galectin-3 | LGALS3 | EOS - SAO |  | ↓ |  | ↑ |  |  |  |  |  |
| Vesicle-associated membrane protein 8 | VAMP8 | All EOS |  | ↑ |  |  |  |  |  |  | platelet secretion and thrombin-PAR signaling |
| Matrix metalloproteinase-10 | MMP10 | EOS-LAA |  | ↓ |  |  |  |  |  |  | atherosclerotic matrix remodeling |
| Lymphatic vessel endothelial hyaluronan receptor 1 | LYVE1 | All EOS |  | ↑ |  |  |  | ↑ |  |  |  |

### Associations with VTE and ischemic stroke-related risk factors

Four of the EOS-associated proteins (*ABO*, *F11*, *GRK5*, and *LGALS3*) were significantly associated with VTE, another prothrombotic condition (Bonferroni p-value threshold = 0.05/19 tests = 0.0026). Two proteins were associated with hypertension (*MLN* and *MEP1B*), two with BMI *(F11* and *LYVE1*), two with T2D (*ABO* and *MEP1B*), and three with LDL-C (*ABO*, *FN1*, and *FCN2*) (**Table 2 and Table S6**).

### Proteins previously reported to predict stroke incidence

We identified six longitudinal studies linking serum or plasma proteins to the risk of ischemic stroke. Proteins were measured on either the OLINK or SOMAScan platform. Characteristics of these studies are provided in **Table S7**. Collectively, these studies reported 76 proteins associated with ischemic stroke incidence. Instruments were available from our study for 32 of these 76 proteins, allowing us to estimate their causal associations with ischemic stroke. The IVW causal estimates are shown in **Table S7**. Of these 32, four were causally associated with EOS in our MR analysis at p < 0.05: ADGRG1 (OR = 0.93; 0.89-0.97; p = 0.002), *CCL7* (OR = 1.16; 1.04-1.28; p = 0.005), *PCSK9* (OR = 1.14; 1.05-1.23; p = 0.001), *PLA2G1B* (OR = 0.58; 0.36-0.92; p = 0.022), and *TFRC* (OR = 0.93; 0.90-0.97; p < 0.001).

### Drug-gene interactions

Using Mendelian randomization, we identified multiple protein-coding genes whose genetically predicted protein levels were causally associated with ischemic early-onset stroke (EOS). We then queried the Drug–Gene Interaction Database (DGIdb) to systematically map known drug–gene interactions to these MR-prioritized genes, yielding actionable candidates concentrated in thrombo-inflammatory pathways (e.g., *F11*, *F7*, *BTD*). Among these, *F11* (Factor XI) showed the most compelling repurposing signal: DGIdb mapped abelacimab (a Factor XI/FXIa inhibitor) to *F11* with a high interaction score, and abelacimab is already in advanced clinical development explicitly for stroke/systemic embolism prevention, supporting *F11* inhibition as a biologically and translationally aligned strategy for ischemic EOS prevention. DGIdb also mapped additional Factor XI–directed agents (e.g., osocimab, milvexian) to *F11*, reinforcing Factor XI/FXIa as a druggable node for antithrombotic intervention with potential relevance to EOS.

A second mechanistically concordant cluster involved complement/contact-system regulation: human C1-esterase inhibitor (C1-INH) was mapped to *F11*, and preclinical ischemia–reperfusion studies have demonstrated that C1-INH can reduce infarct burden and improve functional outcomes through combined anti-inflammatory and antithrombotic effects, consistent with thromboinflammatory contributions to ischemic injury. In contrast, DGIdb hits at *F7* (Factor VII) included recombinant activated factor VIIa and other procoagulant agents; given their mechanism and observed increases in arterial thromboembolic events in clinical testing, these were judged contra-indicated for ischemic EOS prevention. Finally, DGIdb mapped aspirin/salicylic acid to *BTD* and, consistent with established clinical practice, antiplatelet therapy remains a pragmatic, guideline-supported comparator and prevention strategy for non-cardioembolic ischemic stroke, providing a clinically grounded benchmark for evaluating novel repurposing candidates emerging from MR.

## Discussion

Determinants of EOS differ in important ways from those of late-onset stroke, including greater heritability for EOS,^25–27^ differences in the distribution of etiological subtypes, and age-dependent effects of some conventional risk factors, such as BMI.^37^ Using cis-acting genetic variants to strengthen causal inference and improve biological interpretability, we identified 19 proteins associated with EOS or one of its major subtypes. These proteins clustered within four interconnected biological domains: coagulation, platelet adhesion and activation, vascular/extracellular matrix remodeling, and inflammation. The principal novelty of this study lies in applying a proteome-wide Mendelian randomization framework to the largest available genetic study of circulating proteins in EOS, a phenotype whose genetic architecture differs from ischemic stroke occurring later in life.

The strongest causal signals localized to proteins involved in coagulation, most notably ABO, F11, and F7. The Early Onset Stroke Consortium previously established *ABO* as the most significant genome-wide locus for ischemic stroke, with substantially larger effects in EOS than in LOS.^29^ This association is thought to be mediated primarily through ABO-dependent variation in circulating von Willebrand factor (VWF) and factor VIII levels, as well as effects on the molecular properties of these proteins, thereby promoting thrombosis, particularly among individuals with non-O blood groups. *ABO* has also been repeatedly associated with VTE, suggesting that it influences a shared thrombotic susceptibility that extends across both arterial and venous vascular beds. The stronger association of *ABO* with EOS than LOS observed in both the original EOSC GWAS and our proteogenomic analyses suggests that inherited prothrombotic mechanisms may be particularly important in stroke occurring at younger ages. Although *ABO* itself is not directly druggable, its biologic effects converge on established thrombotic pathways involving VWF, factor VIII, platelet adhesion, and coagulation activation, reinforcing the central role of thrombo-inflammatory mechanisms in EOS pathogenesis and further supporting investigation of downstream coagulation pathways as therapeutic targets for stroke prevention.

The overlap between EOS-associated proteins and VTE should not be interpreted as evidence that VTE occurring after stroke contributes to the observed associations. Mendelian randomization evaluates the effects of inherited genetic determinants of protein levels and disease susceptibility and is therefore largely resistant to reverse causation. Accordingly, these shared associations likely reflect common biologic pathways underlying susceptibility to both arterial and venous thrombosis. This interpretation is particularly compelling for *ABO* and *F11*, which are established determinants of thrombotic disease across vascular territories. F11 and F7 represent key components of the intrinsic and extrinsic coagulation pathways, respectively. Factor XI amplifies thrombin generation through the contact activation pathway, whereas factor VII initiates coagulation following endothelial injury through tissue factor binding. Prior Mendelian randomization studies have reported that higher factor XI activity increases ischemic stroke risk, particularly cardioembolic stroke,^38^ while combined GWAS and MR analyses support a causal contribution of factor VII activity to ischemic stroke susceptibility.^39^ These findings reinforce a growing body of evidence indicating that coagulation biology plays a central role in EOS.

The therapeutic implications of these observations are noteworthy. Factor XI has emerged as an attractive antithrombotic target because inhibition of the intrinsic pathway may reduce thrombosis while preserving much of normal hemostatic function. Several factor XI inhibitors, including abelacimab, osocimab, milvexian, and asundexian, are approved or in advanced clinical development. ^40^ Recent clinical trial evidence further supports this strategy; the OCEANIC-STROKE trial reported that asundexian reduced recurrent ischemic stroke and major cardiovascular events among patients with prior non-cardioembolic stroke.^41^ In contrast, therapeutic inhibition of factor VII would be undesirable given its central role in physiological hemostasis. Additional agents with both anti-inflammatory and antithrombotic properties, including C1 esterase inhibitor and aspirin, further highlight the extensive biologic crosstalk among coagulation, platelet activation, and inflammatory pathways.

A second cluster of proteins implicates platelet adhesion, secretion, and activation. VAMP8 is a key regulator of platelet granule release and contributes to amplification of platelet activation and stable thrombus formation.^42^ GRK5 modulates G-protein-coupled receptor signaling, including thrombin-mediated protease-activated receptor (PAR) pathways that influence platelet responsiveness and arterial thrombosis.^43,44^ Notably, GRK5 variants overlap with ischemic stroke susceptibility loci identified in in GIGASTROKE,^45^ providing independent genetic support for its role in cerebrovascular disease.

Our findings also identify *ITGA6* as a biologically compelling candidate within this platelet pathway. *ITGA6* encodes the α6 integrin subunit, which pairs with β1 integrin to form the α6β1 laminin receptor expressed on platelets. This receptor mediates platelet adhesion to laminins exposed within the vascular basement membrane following endothelial injury and contributes to platelet activation and thrombus formation under arterial flow conditions.^46^ Although *ITGA6* has not yet emerged as an established ischemic stroke locus in large-scale GWAS, its role in platelet adhesion and arterial thrombosis directly links it to mechanisms underlying ischemic stroke. Notably, ITGA6 was among the proteins demonstrating a significantly larger causal effect for EOS than for LOS, suggesting that platelet adhesion pathways may be particularly relevant to stroke occurring at younger ages.

A third biologic domain involves vascular remodeling and extracellular matrix (ECM) turnover. *MMP10* and *FN1* point toward pathways governing matrix degradation, fibrous cap stability, and atherosclerotic plaque remodeling. Matrix metalloproteinases contribute to degradation of extracellular matrix components, while fibronectin plays a central role in ECM organization, cellular adhesion, and vascular repair.^47,48^ Perturbations in these pathways may influence plaque progression, plaque instability, and the likelihood of plaque-related thromboembolic events, particularly in large artery atherosclerotic stroke.

*EHBP1* broadens this vascular-remodeling axis through a distinct mechanism involving intracellular trafficking. Although not traditionally viewed as an ECM or stroke gene, *EHBP1* encodes an endosomal trafficking adaptor that regulates vesicular transport and links endosomes to the actin cytoskeleton. Emerging evidence suggests that *EHBP1* influences pathways relevant to tissue remodeling and fibrosis, while recent mechanistic studies demonstrate that EHBP1 regulates sortilin-PCSK9-LDL receptor trafficking, thereby affecting cholesterol handling and potentially atherosclerotic disease progression.^49^ Consistent with this biology, *EHBP1* demonstrated a significantly larger causal effect for EOS than for LOS in our analyses. Although additional validation is required, these observations suggest that *EHBP1* may represent a novel susceptibility factor linking vascular remodeling, lipid metabolism, and cerebrovascular risk.

The fourth major biologic theme emerging from our findings is inflammation. *FCN2* encodes ficolin-2, a pattern-recognition molecule that activates the lectin complement pathway. Increasing evidence implicates complement activation in thrombo-inflammation, vascular injury, and ischemia-reperfusion damage following cerebral ischemia.^50^ Our findings therefore support a role for innate immune activation in EOS pathogenesis and highlight lectin complement signaling as a potentially important contributor to cerebrovascular risk.

*LGALS3* encodes galectin-3, a multifunctional lectin involved in macrophage activation, fibrosis, immune regulation, and sterile inflammation. Although a direct causal role in stroke initiation remains uncertain, galectin-3 has consistently been associated with stroke outcomes, vascular inflammation, and cardiovascular disease.^51^ Interestingly, we observed that higher genetically predicted LGALS3 levels were associated with lower EOS risk but increased VTE risk, suggesting potentially divergent effects across arterial and venous thrombotic phenotypes. These findings underscore the complexity of inflammatory pathways in vascular disease and warrant further investigation into context-specific effects of galectin-3 biology.

Cross-trait analyses further supported the biological relevance of several prioritized proteins. Four proteins (ABO, FN1, ITGA6, and EHBP1) exhibited significantly larger effects in EOS than LOS, whereas ABO, F11, GRK5, and LGALS3 were also associated with VTE. These observations suggest that EOS shares important biological mechanisms with other thrombotic and vascular phenotypes while retaining distinct age-related susceptibility pathways. Collectively, the findings support the view that multiple proteins influence EOS through mechanisms that overlap with, but are not identical to, those underlying conventional cardiovascular risk factors.

We also examined proteins previously associated with ischemic stroke incidence in large epidemiologic studies. Of 76 reported proteins, 32 were represented in our proteomic dataset, and four (*ADGR1*, *CCL17*, *PCSK9*, and *TFRC*) were associated with EOS at p ≤ 0.005. Failure to replicate the remaining proteins may reflect limited statistical power, non-causal epidemiologic associations, differences in outcome definition, or context-specific biological effects. Among these proteins, PCSK9 is particularly noteworthy because it promotes atherosclerosis through enhanced degradation of the LDL receptor and consequent elevation of LDL cholesterol levels. Supporting a causal role, a large UK Biobank phenome-wide association study demonstrated that *PCSK9* loss-of-function variants were associated with substantially lower ischemic stroke risk (OR ≈ 0.61).^52^ Notably, this association was observed despite *PCSK9* not emerging as a genome-wide significant locus in MEGASTROKE, suggesting that genetically mediated modulation of *PCSK9* may influence stroke risk through established lipid pathways that are not readily detected in conventional stroke GWAS.

Our study has several notable strengths, including its focus on EOS, use of the largest currently available EOS genetic resource, and restriction of instrumental variables to cis-acting variants, which improves biological interpretability and reduces susceptibility to horizontal pleiotropy. However, two limitations should be acknowledged. First, we interrogated proteins represented on cardiovascular- and inflammation-focused proteomic panels. Although this hypothesis-driven approach likely enriched for biologically relevant targets, it reduced the overall proteomic coverage and may have limited the discovery of previously unrecognized pathways. Second, participants were exclusively of European ancestry, reflecting both the composition of the UK Biobank proteomic resource and the Early Onset Stroke Consortium. Extension of these analyses to more diverse populations will be essential to assess generalizability, identify ancestry-specific mechanisms, and improve fine-mapping resolution. Finally, we acknowledge that individual-level clinical data were unavailable for sub-analyses because the analyses used summary GWAS statistics from EOSC.

In conclusion, our integrative cis-MR proteogenomic analysis identifies four interconnected biological domains underlying EOS susceptibility: coagulation (ABO, F7, F11), platelet adhesion and activation (ITGA6, VAMP8, GRK5), vascular and extracellular matrix remodeling (FN1, MMP10, EHBP1), and innate inflammatory signaling (FCN2, LGALS3). These findings reinforce ABO as a major locus with amplified effects in early life while nominating ITGA6, EHBP1, VAMP8, GRK5, and LGALS3 as biologically plausible candidates for subtype-specific risk stratification and therapeutic targeting. Collectively, these results provide a focused set of proteins for replication, functional validation, and translational investigation aimed at improving precision prevention and treatment strategies for early-onset ischemic stroke.

**Figure 1:**
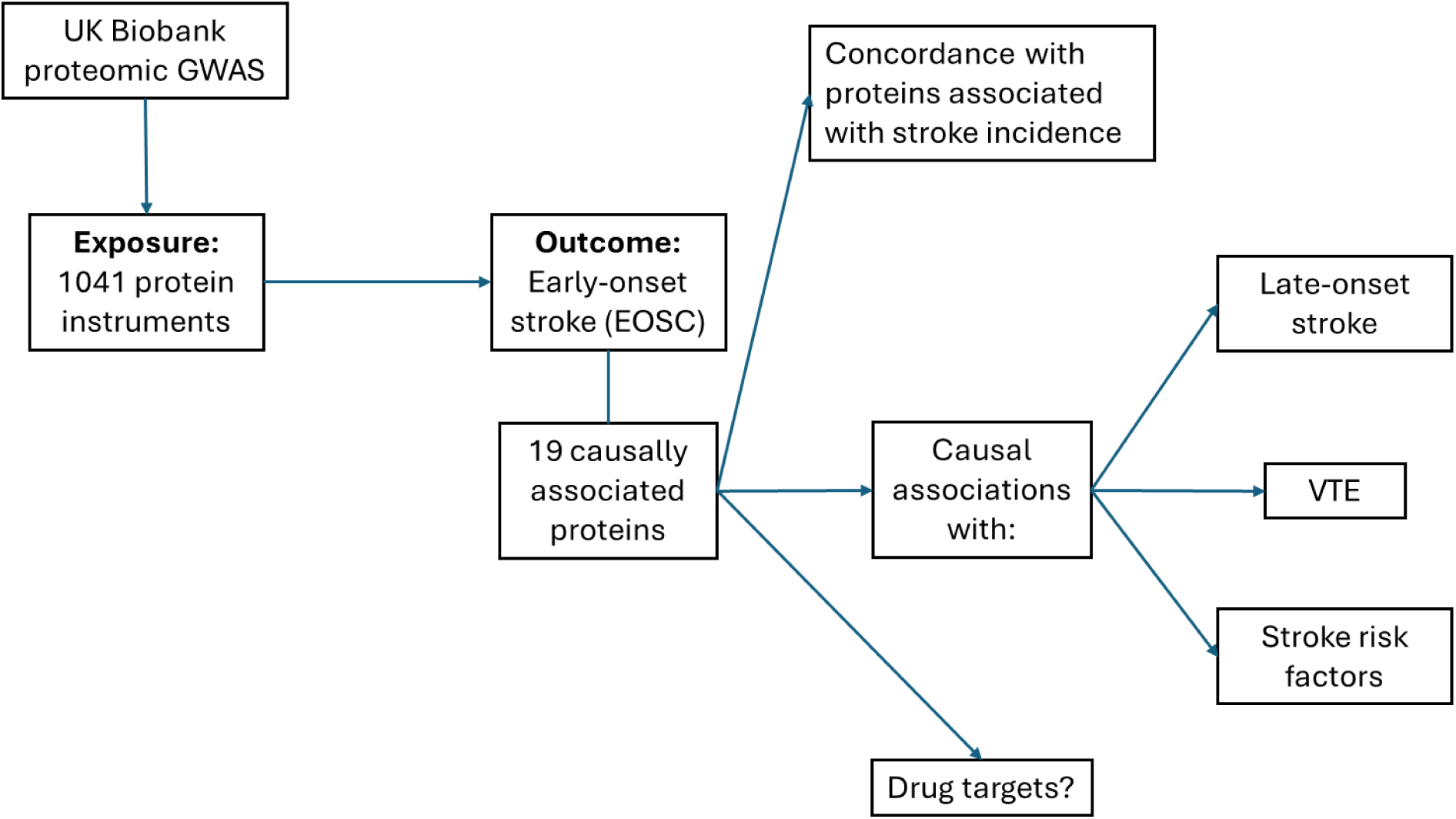
Study design: Mendelian randomization analysis of plasma protein levels on early onset stroke and follow-up analyses of 19 associated proteins. Abbreviations: VTE, venous thromboembolism

**Figure 2.**
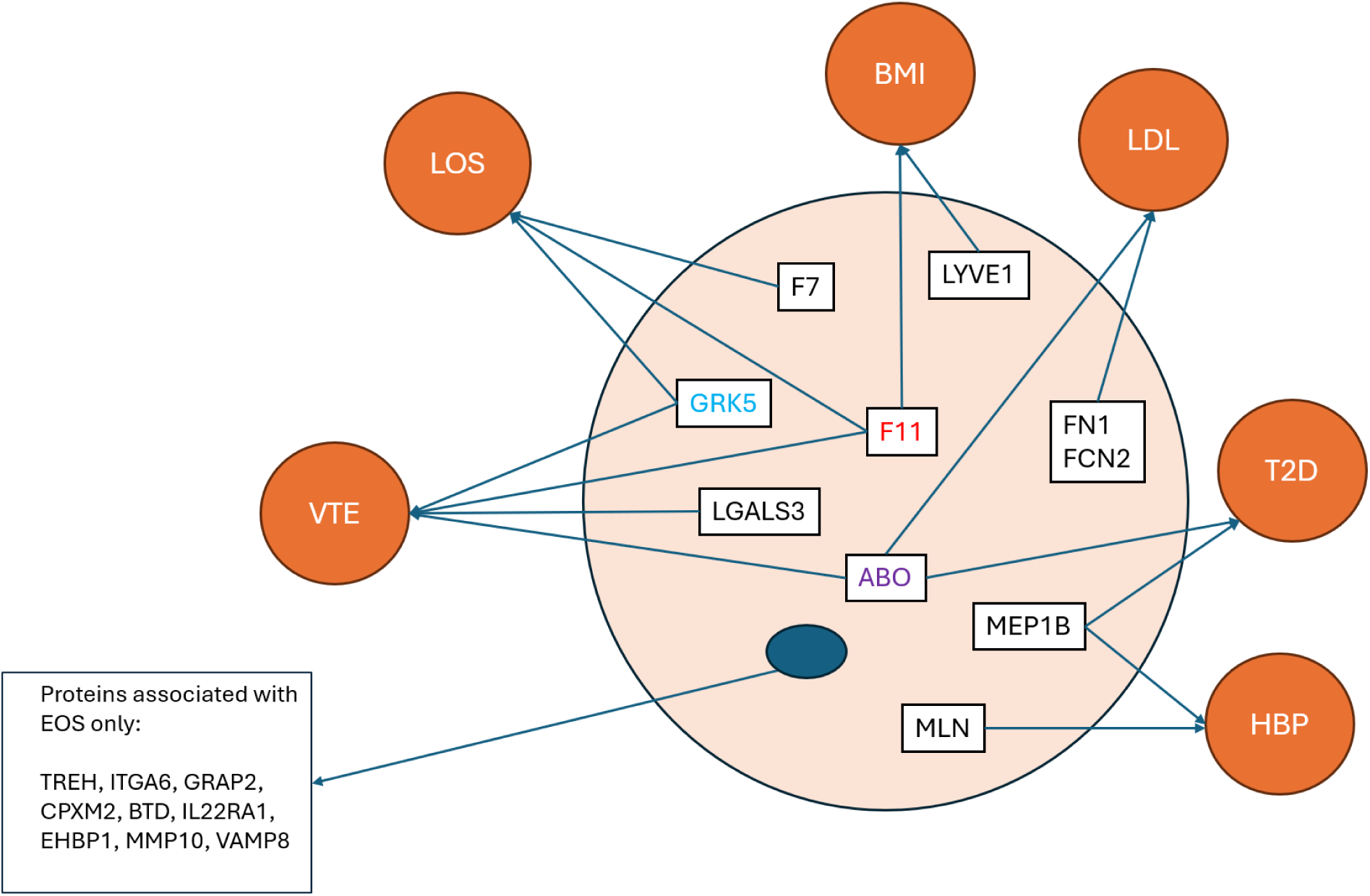
19 EOS-associated proteins and their causal associations with LOS, VTE, and stroke-related risk factors Abbreviations: EOS, early-onset ischemic stroke; LOS, late-onset stroke; VTE, venous thromboembolism; HBP, hypertension; BMI, body mass index; T2D, type 2 diabetes; LDL-C, low-density lipoprotein cholesterol.

## Acknowledgments, Sources of Funding, & Disclosures

**Sources of Funding**. This work was supported by NIH grants R01 NS100178, R01 NS105150, and R01 NS114045 and by Veteran’s Administration grant BX004672. Dr. Xu was supported by the AHA (Grant 19CDA34760258).

**Disclosures:** None.

## Non-standard Abbreviations and Acronyms

MR: Mendelian Randomization
EOS: Early Onset Stroke
LOS: Late Onset Stroke
EOSC: Early Onset Stroke Consortium
SiGN: Stroke Genetics Network
TOAST: Trial of ORG 10172 in Acute Stroke Treatment
CE: Cardioembolic Stroke
LAA: Large Artery Atherosclerosis
SAO: Small Artery Occlusion
OTHER: Stroke of other determined cause
UNDETER: Stroke of undetermined cause
IVW: Inverse Variance Weighted Method
VTE: Venous thromboembolism

